# Seropositivity to Dengue virus (DENV) in three neighborhoods in the periphery of a city with a recent history of outbreaks in Argentina: what can we learn from unreported cases?

**DOI:** 10.1101/2024.07.02.24309829

**Authors:** Diego Antonio Mendicino, Tamara Ricardo, Maximiliano Ariel Cristaldi, Mariana Maglianese, Gastón Guzmán, Sebastián Claussen, Romina Guadalupe Chiaraviglio, Christian Alberto Ávalos, María Andrea Previtali

## Abstract

**Background:** Argentina has experienced several dengue outbreaks since its reemergence in 1998, but cases are underreported due to asymptomatic infections and inadequate access to the healthcare system, particularly in marginalized neighborhoods.

**Methods:** Between December 2019 and March 2020, we assessed seroprevalence of DENV in three neighborhoods in the periphery of the city of Santa Fe. Serum samples were obtained from one adult per household and analyzed by ELISA for DENV IgG antibodies. Multivariate logistic regression models were used to analyze seropositivity by demographic, socio-economic and environmental variables.

**Results:** From 184 participants,overall seroprevalence was 15.2% (95% CI: 10.7; 21.1%). Proximity to a vacant lot decreased the likelihood of seropositivity by 65% (OR: 0.35; 95% CI: 0.15; 0.80). This aligns with studies suggesting vegetation cover can reduce vector abundance and that DENV transmission is higher in densely populated urban areas. Surprisingly, knowledge of disease symptoms and transmission was not linked to lower seropositivity, echoing findings from other studies.

**Conclusions:** These findings contribute to building consensus on the factors that increase DENV infection and will be valuable in designing public health interventions in vulnerable neighborhoods during dengue outbreaks.

## 1. Introduction

The emergence or re-emergence of infectious diseases (EID) stands as one of the primary consequences of climate change, posing a growing concern in public health, particularly in the aftermath of the COVID-19 pandemic [1,2]. Disease reservoirs and vectors respond to these environmental changes by migrating to more favorable areas, expanding their geographic distribution, or modifying their reproductive behavior [1,2].

Dengue fever, caused by dengue virus (DENV serotypes 1-4) and transmitted by the bites of *Aedes aegypti* and *Ae. albopictus* mosquitoes, is a vector-borne EID that has spread to urban environments in subtropical and temperate regions of the world [3,4]. In these settings, *Ae. aegyptis* and *Ae. albopictus* can breed inside water containers stored within houses and in the peridomiciliary environment [3,4]. The disease dynamics presents a marked seasonality, with an increase in cases during warm and humid months where *Aedes* mosquitoes have the higher activity [3].

Dengue re-emerged in Argentina in 1998 and has registered periodic outbreaks in the northern region of the country since then [5,6]. Dengue is a disease of mandatory notification in Argentina [7]. However, in many cases, the infection is asymptomatic or oligosymptomatic, and sometimes symptoms are confused with other infectious diseases, leading to biases in the record of the community circulation [4,7]. Inadequate access to healthcare in marginalized sectors of society, combined with geographic accessibility to healthcare facilities, can determine the possibilities of getting timely testing and diagnosis [8,9].

The province of Santa Fe, located in northeastern Argentina, presents suitable conditions for the proliferation of *Aedes* mosquitoes, such as heavy rainfall and flooding, heat waves, and unplanned urban expansion with an increasing number of informal settlements and urban slums [1,2,6]. Santa Fe has experienced dengue outbreaks in the years 2009, 2016, 2019, and 2020 [5,6] and is currently experiencing a severe outbreak, with 51,249 confirmed cases reported between epidemiological weeks 1 and 19 of 2024 [10]. The 2020 outbreak included 4,670 confirmed cases, representing an incidence of 81.6 cases per 10,000 inhabitants (95% CI: 79.3-84.0 cases per 10,000 inhabitants), with a peak of cases occurring between epidemiological weeks 7 and 22 (Fig. S1) [6,10]. This outbreak overlapped with the first records of COVID-19 cases in the country, representing a double challenge for health services [11]. Although several studies addressed the epidemiology of dengue in Argentina in the period 2009-2020 [6,9,11–15], most of them analyzed secondary data and only one of them analyzed data from Santa Fe [5]. The objective of our study was to determine seroprevalence of DENV in three neighborhoods in the periphery of the city of Santa Fe, Argentina.

## 2. Materials and methods

### 2.1. Study design and setting

The city of Santa Fe, the capital of the province of Santa Fe, Argentina, is surrounded by rivers on the eastern, western, and southern sides, with over 70% of its total surface consisting of rivers, lagoons, and wetlands (Fig. 1). Santa Fe has a humid subtropical climate, with mean annual temperatures of 18-20°C and mean annual precipitation between 1000-1200 mm [16]. Socioeconomic conditions vary widely within the city limits, with a higher concentration of urban slums and informal settlements observed in the riverside areas [17].

**Figure 1.**
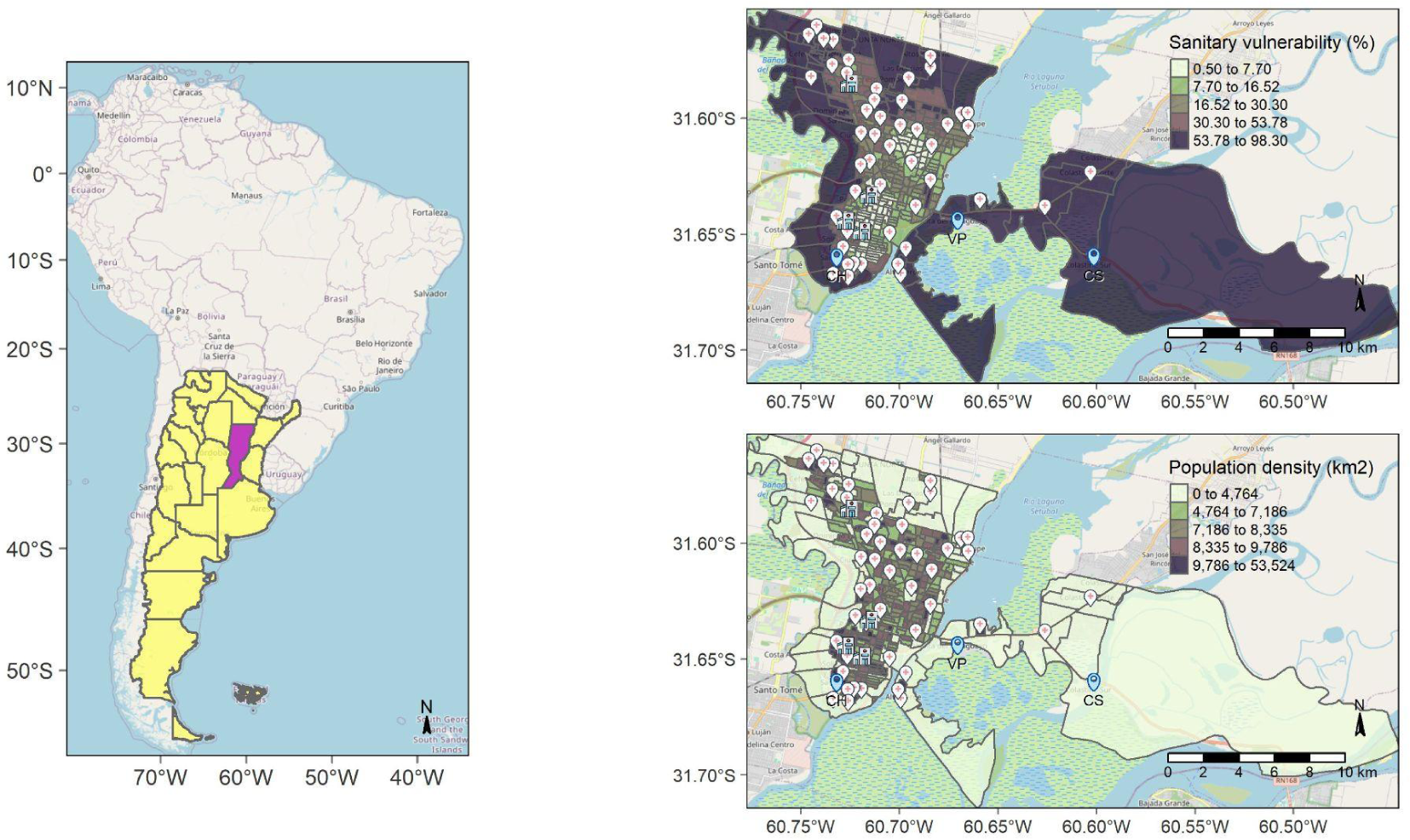
Map of study area. The left panel shows the location of Argentina in South America and the location of Santa Fe in Argentina. The right panels show a zoomed view of the city of Santa Fe, with census tracts colored by sanitary vulnerability (top right) or by population density (bottom right). Map pins indicate the location of the sampled neighborhoods, public health units and public hospitals. Spatial layers from OpenStreetMap® and POBLACIONES [18]; map pin icons from Freepik. An interactive version of this map is available at https://issengard83.github.io/KAP_dengue_2020/

Between December 2019 and March 2020, we conducted a cross-sectional survey targeting residents in three neighborhoods of Santa Fe: Chalet (CH), Colastiné Sur (CS), and La Vuelta del Paraguayo (VP) (Fig. 1). The three neighborhoods present high percentages of unsatisfied basic needs and health vulnerability [19] (Fig. 1), as well as deficiencies in basic infrastructure such as: lack of sewage system, unpaved or semi-paved roads, irregular garbage collection, and, in the case of CS, lack of access to piped water [18]. These neighborhoods also differ in whether there is a healthcare unit on site and the proximity to the public hospitals (Fig. 1). By encompassing three different settings, we intended to identify demographic, socio-economic and environmental conditions that could be associated with the seropositivity to DENV.

### 2.2. Participants

Sample size was estimated using R version 4.4.0 [19] with the *samplingbook* package [20], incorporating a correction for finite population. Population sizes were determined based on the number of households in the census tract corresponding to each sampled neighborhood, obtained from the 2010 National Census [21]: 277 households for CH, 314 for CS, and 99 for VP. Assuming an expected seroprevalence of 50% and a precision of 10%, the calculated sample sizes were as follows: 72 households for CH, 74 for CS, and 49 for VP.

Selected participants must be at least 18 years old and have resided in the neighborhood for one year or more. Exclusions from the study were individuals who do not permanently reside in the neighborhood, those who declined to provide a serum sample for dengue testing, and those with known previous dengue infections. In order to prevent pseudoreplication, we only sampled one person per household.

### 2.3. Data collection tool

Participants were requested to respond to an interviewer-administered questionnaire to provide information on the following sociodemographic variables: gender (male, female, other), age (years), education (illiterate, incomplete primary school, primary school, incomplete high school, high school, incomplete tertiary or university, tertiary or university), occupation (homemaker, student, retired, unemployed, underemployed, self-employed, public employee, private employee). The level of urbanization of the study sites was assessed using the following variables: accumulation of rainwater or flooding within the 50 meters around the home (yes, no), time since the last accumulation of rainwater/flooding (open-ended question), area reached by the flood water (street, front/backyard, inside the house), time taken for the water to recede (hours, days, weeks), accumulation of water containers in the household (yes, no), source of drinking water (water pipes, spring or well, tank truck), use of environmental water for cleaning or recreation (yes, no), method of disposing of garbage (garbage truck, burning, accumulation in the backyard, other/s), dump sites within the 50 meters around the home (yes, no).

Additionally, participants were asked yes/no questions about their awareness of dengue, if they knew someone who had had dengue, if that person lived in the neighborhood, and if that person lived in their household. Open-ended questions were included to allow for multiple answers on identification of dengue symptoms (does not know/no response, fever, headache, myalgia, malaise, skin rash, fatigue, diarrhea, vomits, bleeding), ways of transmission (does not know/no response, travel to endemic areas, mosquito bites, storage of water containers, person-to-person, sexual intercourse), presence of symptoms of febrile illness in the previous six months (none, fever, diarrhea and/or vomiting, skin rash, headache, myalgia, malaise), and actions taken in case of symptoms of febrile illness (none, self-medication, healer/shaman, health center).

The research team also collected information through direct observation on the following variables: street type (paved road, semi-paved road, dirt road, sand), proximity to gutters and ditches (yes, no), distance to gutters and ditches (less than 10m, 10-25m, 25-50m), proximity to dumpyards (yes, no), distance to dumpyards (less than 10m, 10-25m, 25-50m), proximity to vacant lots (yes, no), distance to vacant lots (less than 10m, 10-25m, 25-50m), proximity to water bodies (yes, no), distance to the water bodies (less than 10m, 10-25m, 25-50m), house floor impermeability (yes, no), house roof impermeability (yes, no). After the completion of the survey, an informative flyer with the most common symptoms of dengue, modes of transmission and preventive actions was given to each participant and explained.

### 2.4. Laboratory analysis

Approximately 5 ml of blood was drawn by laboratory technicians from each participant through venipuncture, using sterile needles and syringes and collected in sterile tubes containing coagulation activator and separator gel (BD Vacutainer). After allowing the clot to form for 15 minutes at room temperature, the blood samples and disposable material were transported to the laboratory. Samples were centrifuged at 3500 revolutions per minute (rpm) for 10 minutes. During the transfer to the laboratory, samples were kept refrigerated at 4-8 °C in the primary tubes, then stored at 2-8°C, and processed within 5 days. Biosafety standards were respected throughout the process.

All samples were analyzed by Enzyme-linked immunosorbent assay (ELISA), which uses inactivated and purified Dengue virus types 1-4 to detect Immunoglobulin G specific antibodies (Dengue Virus IgG DxSelect Focus Diagnostics). The manufacturer’s instructions were followed for processing the ELISA tests and a Micropar Washer washer and a Mindray MR-96A reader were used during the procedures. We calculated the cut off in each batch of samples. The specificity and sensitivity of the kit were 0.93 (95% IC: 0.90-0.96) and 0.96 (95% IC: 0.89-0.99), respectively. The results were handed out to each participant as printed reports together with a flyer with preventive measures, along with in person clarifications and advice to prevent the disease.

### 2.5. Data processing

Age groups of the participants were categorized based on the age quartiles of the sample (18-31 years, 32-43 years, 44-58 years, 59+ years). Education levels were recategorized into three groups (illiterate/incomplete primary school, primary school/incomplete high school, high school and higher), and occupations were re-categorized into four groups (homemaker/student, retired/pensioner, unemployed/underemployed, employed) based on the frequency of responses. Street type was collapsed into three categories: paved or semi-paved road, dirt road, or sand. The variables “distance to water bodies”, “distance to roadside channels and ditches” and “distance to dump-yards” were re-categorized into: <25 meters, 25-50 meters and >50 meters.

We created a variable to indicate the accumulation of rainwater or flooding within the 50 meters around the home in the previous 30 days (yes, no), derived from the responses to the question about the time since the last accumulation of rainwater or flooding. Additionally, we created a variable to document whether the participant stores rainwater, based on the comments provided in the section about the source of water for drinking and cleaning. The methods of garbage disposal were categorized into four yes/no variables: garbage truck, burning, accumulation in the backyard, and throwing at the river.

We considered participants to be aware of dengue symptoms if they selected fever along with any of the following symptoms: headache, muscle pain, retro-orbital pain, rash, nausea and vomiting, diarrhea, or hemorrhagic manifestations [7]. Similarly, we determined that participants were aware of the modes of transmission of dengue if they selected mosquito bites, keeping water containers inside or outside the house, or traveling to endemic areas [7]. We created a variable indicating whether participants had a previous dengue infection, using information from the comments section of the question asking if they knew someone who had had dengue.

### 2.6. Data analysis

We entered data into Microsoft Access® and performed cleaning, processing, and analysis using R software, version 4.4.0 [19]. The frequencies of sociodemographic and household characteristics by neighborhood were compared using Pearson’s Chi-squared test or Fisher’s exact test, while the median (IQR) of continuous variables was compared using the Wilcoxon rank-sum test or Kruskal-Wallis rank-sum test using the package *gtsummary* [22].

The same approach was used to assess associations between seropositivity to dengue virus and the following variables: neighborhood (CH, CS, VP), gender, age (years), age group, education level, occupation, street type, proximity to roadside channels and ditches, distance to roadside channels and ditches, proximity to dump-yards, distance to dump-yards, proximity to water bodies, distance to water bodies, proximity to vacant lots, accumulation of rainwater or flooding within the 50 meters around the home in the previous 30 days, accumulation of water containers, source of drinking water, awareness of dengue symptoms, awareness of ways of transmission, if they knew someone who had had dengue, if the person was someone from the neighborhood, if the person was someone from their household, and presence of symptoms of febrile illness in the previous six months.

The variables with a *p*-value below 0.10 in these association tests were selected as explanatory variables for the fit of a set of multivariate logistic regression models. Based on these results, we tested the following associations: (i) interaction between street type and proximity to vacant lots, controlling for the accumulation of water containers in the front or backyards, (ii) interaction between street type and accumulation of water containers in the front or backyards, controlling by proximity to vacant lots, and (iii) interaction between proximity to vacant lots and accumulation of water containers in the front or backyards, controlling by street type. The models were fitted both with and without including neighborhood as a random intercept, resulting in a list of six candidate models. We did not fit models with higher-order interaction terms to avoid complex statistical relationships, overfitting of the data and convergence problems. The fit of multivariate models were compared based on Akaike Information Criteria (AIC) and the Bayesian Information Criteria (BIC) using the package *performance* [23]. Non-significant explanatory variables were removed from the saturated model with the best performance, using a manual *step-backwards* selection. Inferences were drawn from the final model and interpreted in terms of odds ratios (OR) and its 95% confidence interval (95% CI).

Anonymized datasets and R scripts used for this manuscript are available at: Mendicino, D., Ricardo, T., Cristaldi, M., Maglianesi, M., Guzmán, G., Claussen, S., Chiaraviglio, R. G., AVALOS, C., & Previtali, M. A. (2024). Data for Seropositivity to Dengue virus DENV in three neighborhoods in the periphery of a city with a recent history of outbreaks in Argentina: what can we learn from unreported cases? (version1.0) [Data set]. Zenodo. https://doi.org/10.5281/zenodo.11507563

### 2.7. Ethics statement

All participants were informed about the risks and benefits of blood collection and provided written informed consent for the serological analysis and the questionnaires. Datasets have been anonymized to remove personally identifiable information. The procedures adhered to the ethical standards outlined in the Personal Data Protection Law of Argentina (N° 25326) and the principles of the Declaration of Helsinki, 1964, as revised in 1975, 1983, 1989, 1996, and 2000. This research project was evaluated and received approval from the Ethics and Security Advisory Committee in Research of the School of Biochemistry and Biological Sciences of the Universidad Nacional del Litoral, Santa Fe, Argentina (Act No. 03.17).

## 3. Results

### 3.1. Overview of the survey participants

A total of 272 residents from the three selected neighborhoods were initially considered as eligible for the survey. Of these, 81 were excluded due to refusal to provide a blood sample, three participants were excluded as they resided in the same household as others, and an additional four participants were excluded due to prior dengue infections. Among the remaining 184 participants, 66 were from CH, 79 from CS and 39 from VP.

Of the 184 study participants, 59.8% were female and 40.2% were male, with no significant differences among neighborhoods (Table 1). The median age was 44 years (IQR: 32-56 years), and it was significantly lower in participants from VP (Table 1). The maximum education level achieved also differed significantly among neighborhoods, with CH having a significantly higher percentage of participants who finished high school than CS and VP (Table 1). Regarding occupation, 43.4% of the participants identified themselves as employed, 22% as homemakers or students, 20.3% as retired or pensioners, and 14.3% as unemployed or underemployed, with no significant differences among neighborhoods (Table 1).

**Table 1.** Descriptive statistics of the survey participants socioeconomic characteristics and household features by neighborhood, Santa Fe, Argentina (2019-2020). Significant differences among study neighborhoods are highlighted in bold.

| Characteristic | Total<sup>1</sup> | CH<sup>1</sup> | CS<sup>1</sup> | VP<sup>1</sup> | p-value<sup>2</sup> |
| --- | --- | --- | --- | --- | --- |
| **Gender** |  |  |  |  | 0.8 |
| female | 110 (59.8%) | 38 (57.6%) | 47 (59.5%) | 25 (64.1%) |  |
| male | 74 (40.2%) | 28 (42.4%) | 32 (40.5%) | 14 (35.9%) |  |
| **Age (years)** | 44 (32, 56) | 50 (35, 59) | 45 (32, 58) | 37 (28, 44) | **0.014** |
| **Age group** |  |  |  |  | 0.053 |
| 18-31 years | 45 (24.5%) | 11 (16.7%) | 20 (25.3%) | 14 (35.9%) |  |
| 32-43 years | 46 (25.0%) | 14 (21.2%) | 18 (22.8%) | 14 (35.9%) |  |
| 44-58 years | 52 (28.3%) | 24 (36.4%) | 21 (26.6%) | 7 (17.9%) |  |
| 59+ years | 41 (22.3%) | 17 (25.8%) | 20 (25.3%) | 4 (10.3%) |  |
| **Education** |  |  |  |  | **0.018** |
| illiterate/inc. primary school | 23 (12.5%) | 7 (10.6%) | 9 (11.4%) | 7 (17.9%) |  |
| primary school/inc. high school | 89 (48.4%) | 24 (36.4%) | 41 (51.9%) | 24 (61.5%) |  |
| high school and/or university | 72 (39.1%) | 35 (53.0%) | 29 (36.7%) | 8 (20.5%) |  |
| **Occupation** |  |  |  |  | 0.2 |
| homemaker/student | 40 (22.0%) | 11 (16.9%) | 15 (19.0%) | 14 (36.8%) |  |
| retired/pensioner | 37 (20.3%) | 13 (20.0%) | 20 (25.3%) | 4 (10.5%) |  |
| unemployed/underemployed | 26 (14.3%) | 10 (15.4%) | 10 (12.7%) | 6 (15.8%) |  |
| employed | 79 (43.4%) | 31 (47.7%) | 34 (43.0%) | 14 (36.8%) |  |
| **Street type** |  |  |  |  | **<0.001** |
| paved or semi-paved | 56 (30.9%) | 55 (83.3%) | 1 (1.3%) | 0 (0.0%) |  |
| dirt | 27 (14.9%) | 6 (9.1%) | 5 (6.4%) | 16 (43.2%) |  |
| sand | 98 (54.1%) | 5 (7.6%) | 72 (92.3%) | 21 (56.8%) |  |
| **Prox. to roadside channels and ditches** | 61 (33.2%) | 18 (27.3%) | 29 (36.7%) | 14 (35.9%) | 0.4 |
| **Prox. to dumpyards** | 98 (53.3%) | 33 (50.0%) | 37 (46.8%) | 28 (71.8%) | **0.031** |
| **Prox. to vacant lots** | 116 (63.0%) | 23 (34.8%) | 63 (79.7%) | 30 (76.9%) | **<0.001** |
| **Prox. to water bodies** | 71 (38.6%) | 3 (4.5%) | 40 (50.6%) | 28 (71.8%) | **<0.001** |
| **Floor impermeability** | 167 (93.8%) | 61 (95.3%) | 72 (94.7%) | 34 (89.5%) | 0.5 |
| **Roof impermeability** | 160 (89.9%) | 61 (95.3%) | 70 (92.1%) | 29 (76.3%) | **0.011** |
| **Acc. of rainwater or flooding** | 135 (73.4%) | 32 (48.5%) | 67 (84.8%) | 36 (92.3%) | **<0.001** |
| **Acc. of rainwater or flooding (30d)** | 64 (34.8%) | 9 (13.6%) | 44 (55.7%) | 11 (28.2%) | **<0.001** |
| **Source of drinking water** |  |  |  |  | **<0.001** |
| tank truck/other | 81 (44.0%) | 2 (3.0%) | 79 (100.0%) | 0 (0.0%) |  |
| water pipes | 103 (56.0%) | 64 (97.0%) | 0 (0.0%) | 39 (100.0%) |  |
| **Acc. of water containers** | 100 (54.3%) | 27 (40.9%) | 53 (67.1%) | 20 (51.3%) | **0.006** |
| **Regular garbage collection** | 130 (70.7%) | 61 (92.4%) | 62 (78.5%) | 7 (17.9%) | **<0.001** |
| **Storage of rainwater** | 14 (7.6%) | 1 (1.5%) | 13 (16.5%) | 0 (0.0%) | **<0.001** |
| **Garbage acc. in the backyard** | 21 (11.4%) | 0 (0.0%) | 9 (11.4%) | 12 (30.8%) | **<0.001** |

<sup>1</sup>n (%); Median (IQR); <sup>2</sup>Pearson's Chi-squared test; Kruskal-Wallis rank sum test; Fisher's exact test

The site CH was significantly more urbanized than VP and CS, as reflected by the greater coverage of paved roads and regular garbage collection, the lesser presence of vacant lots and accumulations of rainwater or flooding, and the longer distance to water bodies (Table 1). The storage of water containers in the front or backyards was significantly lower than in the other two sites (Table 1).

The site VP presented significantly higher frequencies of permeable roofs (*p* = 0.011), proximity to dumpyards (*p* = 0.031), and accumulation of garbage in the backyard (*p* < 0.001) than sites CH and CS (Table 1). Almost all participants from site CS were either supplied with drinking water by the tank truck (91%) or used other water sources such as springs, wells, or purchased bottled water (8.5%, Table 1). Participants from CS also reported collecting rainwater for cleaning or gardening significantly more often than participants from CH and VP (Table 1). No significant differences were observed among neighborhoods regarding proximity to roadside channels and ditches or floor impermeability (Table 1).

### 3.2. Seropositivity to dengue virus

Twenty-eight out of 184 participants (15.2%, 95% CI: 10.7; 21.1%) were seropositive for IgG antibodies against the dengue virus. The highest seroprevalence was found in site CH (26.9%, 95% CI: 16.8; 40.3%), followed by site VP (18.2%, 95% CI: 8.6; 34.4%) and CS (11.3%, 95% CI: 5.8; 20.7%). Differences in the seroprevalence among neighborhoods were not statistically significant (*p* = 0.2, Table 2). Seropositivity of DENV did not differ significantly by gender, age, age group, education level, or occupation of the participants (Table 2).

**Table 2.**
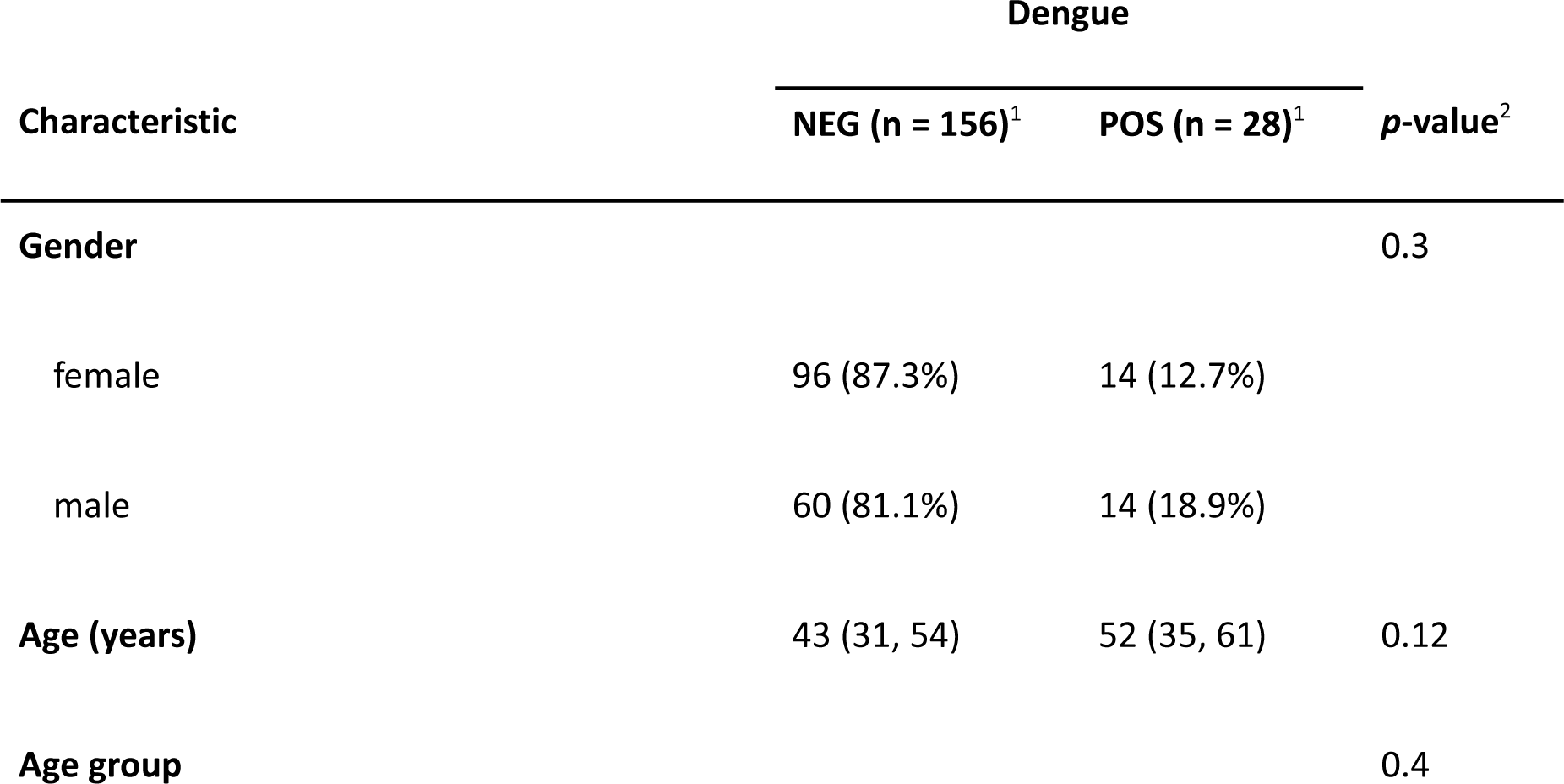
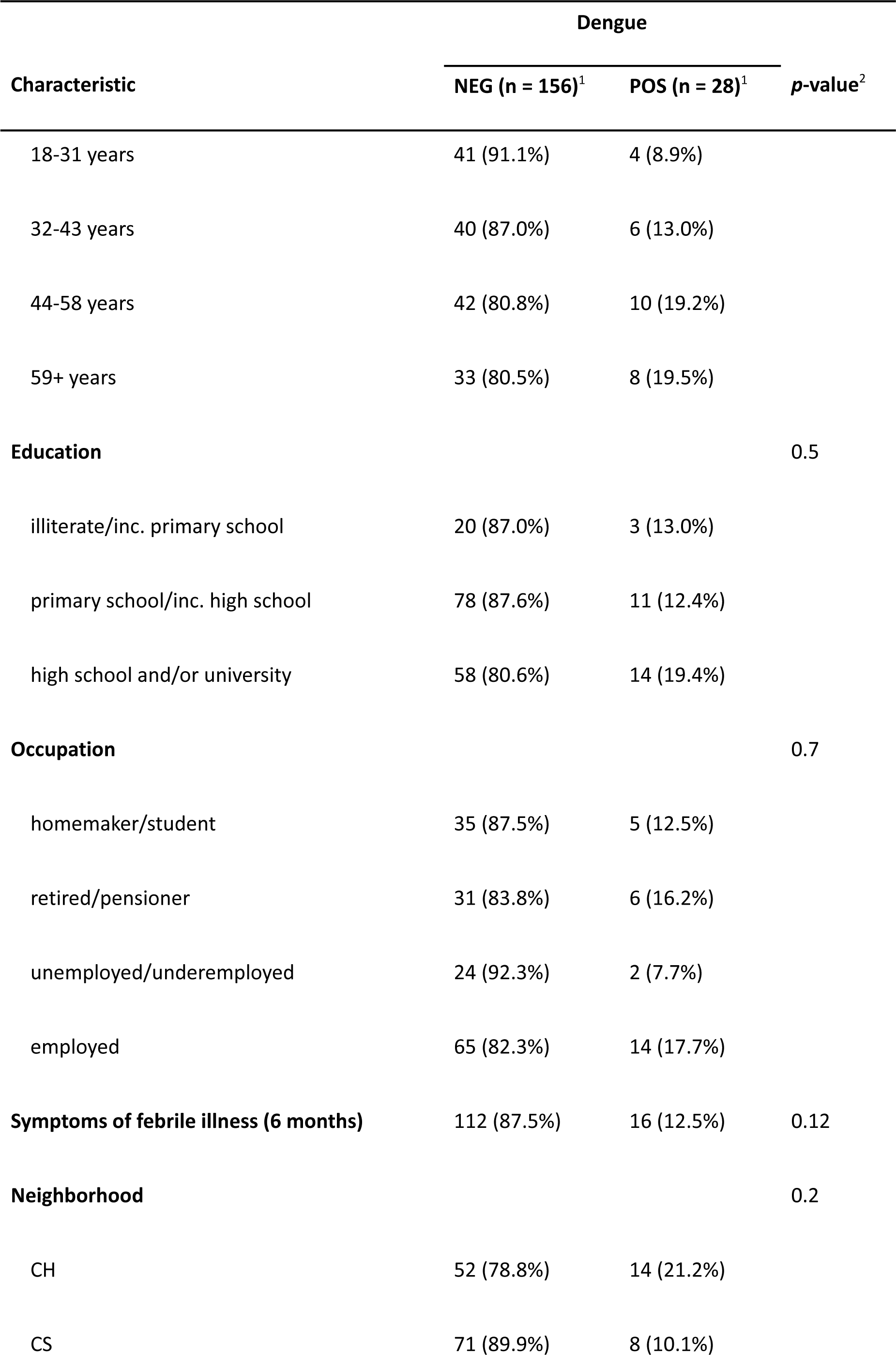
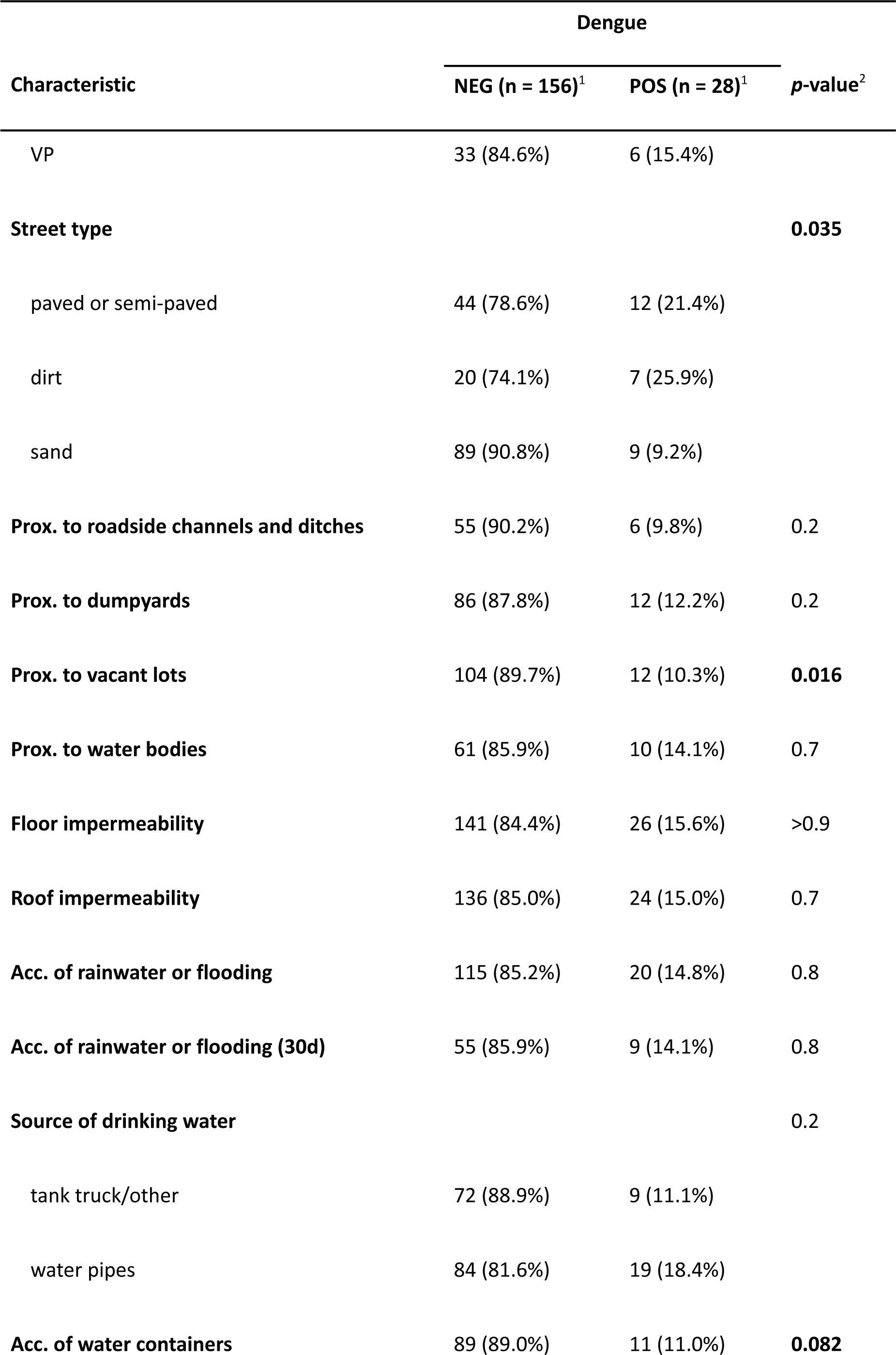
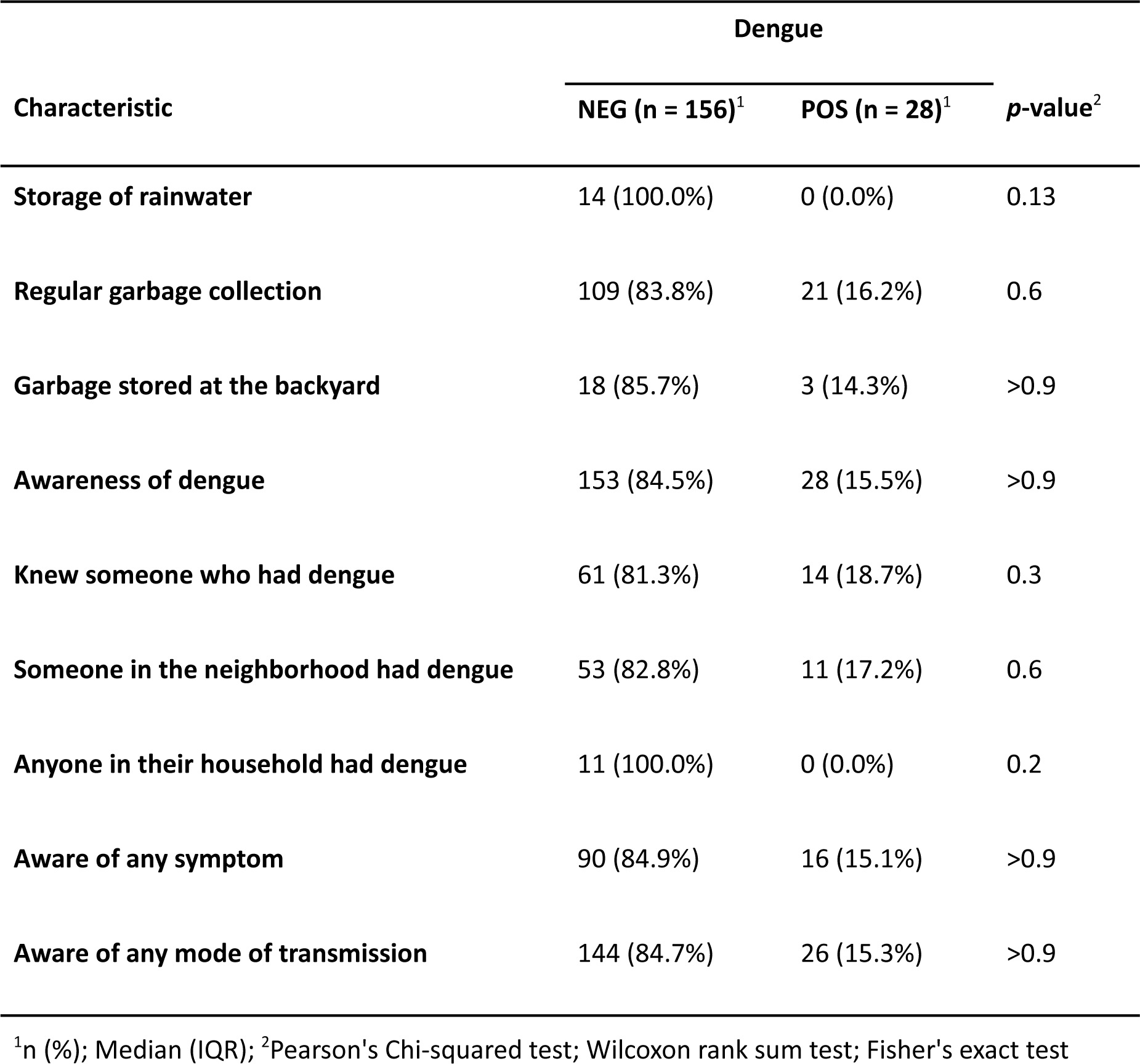
Seropositivity of DENV by sociodemographic and household characteristics of the participants, Santa Fe, Argentina (2019-2020). Variables considered for logistic regression models are highlighted in bold (*p* < 0.1).

| Characteristic | Dengue |  | *p*-value<sup>2</sup> |
| --- | --- | --- | --- |
|  | NEG (n = 156)<sup>1</sup> | POS (n = 28)<sup>1</sup> |  |
| **Gender** |  |  | 0.3 |
| female | 96 (87.3%) | 14 (12.7%) |  |
| male | 60 (81.1%) | 14 (18.9%) |  |
| **Age (years)** | 43 (31, 54) | 52 (35, 61) | 0.12 |
| **Age group** |  |  | 0.4 |

| Characteristic | Dengue |  | p-value<sup>2</sup> |
| --- | --- | --- | --- |
|  | NEG (n = 156)<sup>1</sup> | POS (n = 28)<sup>1</sup> |  |
| 18-31 years | 41 (91.1%) | 4 (8.9%) |  |
| 32-43 years | 40 (87.0%) | 6 (13.0%) |  |
| 44-58 years | 42 (80.8%) | 10 (19.2%) |  |
| 59+ years | 33 (80.5%) | 8 (19.5%) |  |
| **Education** |  |  | 0.5 |
| illiterate/inc. primary school | 20 (87.0%) | 3 (13.0%) |  |
| primary school/inc. high school | 78 (87.6%) | 11 (12.4%) |  |
| high school and/or university | 58 (80.6%) | 14 (19.4%) |  |
| **Occupation** |  |  | 0.7 |
| homemaker/student | 35 (87.5%) | 5 (12.5%) |  |
| retired/pensioner | 31 (83.8%) | 6 (16.2%) |  |
| unemployed/underemployed | 24 (92.3%) | 2 (7.7%) |  |
| employed | 65 (82.3%) | 14 (17.7%) |  |
| **Symptoms of febrile illness (6 months)** | 112 (87.5%) | 16 (12.5%) | 0.12 |
| **Neighborhood** |  |  | 0.2 |
| CH | 52 (78.8%) | 14 (21.2%) |  |
| CS | 71 (89.9%) | 8 (10.1%) |  |
|  | NEG (n = 156)<sup>1</sup> | POS (n = 28)<sup>1</sup> |  |
| VP | 33 (84.6%) | 6 (15.4%) |  |
| **Street type** |  |  | **0.035** |
| paved or semi-paved | 44 (78.6%) | 12 (21.4%) |  |
| dirt | 20 (74.1%) | 7 (25.9%) |  |
| sand | 89 (90.8%) | 9 (9.2%) |  |
| **Prox. to roadside channels and ditches** | 55 (90.2%) | 6 (9.8%) | 0.2 |
| **Prox. to dumpyards** | 86 (87.8%) | 12 (12.2%) | 0.2 |
| **Prox. to vacant lots** | 104 (89.7%) | 12 (10.3%) | **0.016** |
| **Prox. to water bodies** | 61 (85.9%) | 10 (14.1%) | 0.7 |
| **Floor impermeability** | 141 (84.4%) | 26 (15.6%) | >0.9 |
| **Roof impermeability** | 136 (85.0%) | 24 (15.0%) | 0.7 |
| **Acc. of rainwater or flooding** | 115 (85.2%) | 20 (14.8%) | 0.8 |
| **Acc. of rainwater or flooding (30d)** | 55 (85.9%) | 9 (14.1%) | 0.8 |
| **Source of drinking water** |  |  | 0.2 |
| tank truck/other | 72 (88.9%) | 9 (11.1%) |  |
| water pipes | 84 (81.6%) | 19 (18.4%) |  |
| **Acc. of water containers** | 89 (89.0%) | 11 (11.0%) | **0.082** |
|  | NEG (n = 156)<sup>1</sup> | POS (n = 28)<sup>1</sup> |  |
| Storage of rainwater | 14 (100.0%) | 0 (0.0%) | 0.13 |
| Regular garbage collection | 109 (83.8%) | 21 (16.2%) | 0.6 |
| Garbage stored at the backyard | 18 (85.7%) | 3 (14.3%) | >0.9 |
| Awareness of dengue | 153 (84.5%) | 28 (15.5%) | >0.9 |
| Knew someone who had dengue | 61 (81.3%) | 14 (18.7%) | 0.3 |
| Someone in the neighborhood had dengue | 53 (82.8%) | 11 (17.2%) | 0.6 |
| Anyone in their household had dengue | 11 (100.0%) | 0 (0.0%) | 0.2 |
| Aware of any symptom | 90 (84.9%) | 16 (15.1%) | >0.9 |
| Aware of any mode of transmission | 144 (84.7%) | 26 (15.3%) | >0.9 |

<sup>1</sup>n (%); Median (IQR); <sup>2</sup>Pearson's Chi-squared test; Wilcoxon rank sum test; Fisher's exact test

Significant associations were found between seropositivity and street type, and with proximity to vacant lots (Table 2), and marginal associations were observed with storage of water containers (*p* = 0.082, Table 2). No significant associations were detected between seropositivity and any of the other tested variables (Table 2).

The model containing interaction between street type and proximity to vacant lots, controlled by accumulation of water containers in the front or backyards and no random intercept, showed the best performance score (Table 3). After a manual *step-backward* variable selection process only proximity to vacant lots was retained as an explanatory variable. According to this final model, having a vacant lot in the proximity of the domicile (<50m) decreases by 65% the probabilities of seropositivity to dengue (OR: 0.35; 95% CI: 0.15; 0.80), however, this model was able to explain only 3.6% of the observed variability in the data. The model fit met the assumptions for normality and homogeneity of variance of residuals and no outliers were detected.

**Table 3.**
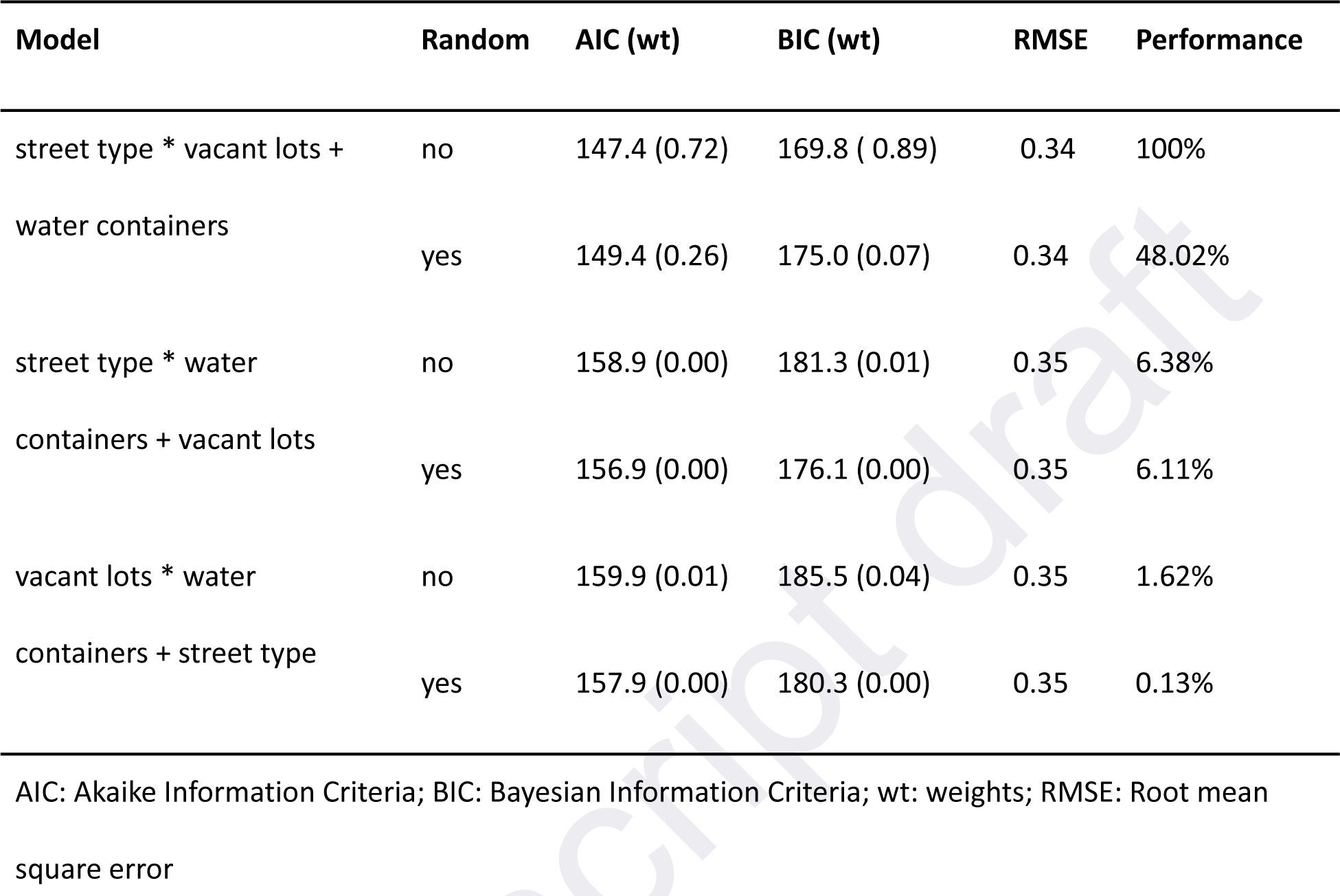
List of candidate multivariate logistic regression models. Random effect: (1|neighborhood). Santa Fe, Argentina (2019-2020).

## 4. Discussion

This is the first field study on dengue seroprevalence and associated risk factors in three neighborhoods of the city of Santa Fe, a city of Argentina with a recent history of DENV outbreaks. Most of the published studies on the incidence of DENV in Argentina were based on cases reported to the Ministry of Health [6,9,11–14]. Therefore, the insights derived from those investigations only pertain to symptomatic cases of dengue reported to the national health surveillance system. Instead, our work involves unreported cases and data on risk factors collected *in situ*.

It has been documented that asymptomatic individuals infected with DENV can transmit the virus to mosquitoes and may be significantly more infectious than symptomatic ones [24,25], posing a risk to further spreading the outbreak within the city and potentially disseminating it to other cities as well. Additionally, people with a secondary DENV infection with a different serotype, even if the primary infection was asymptomatic or oligosymptomatic, are at increased risk of dengue hemorrhagic fever, a potentially fatal clinical form [26].

This study highlights the importance of population screening for estimating dengue seroprevalence, as it provides essential data on risk distribution to plan primary and secondary prevention strategies and to define priorities in healthcare resource allocation. The presence of asymptomatic or unreported dengue cases should be considered when defining priorities for implementing vaccination campaigns [4].

The observed seroprevalence (15.2%, 95% CI: 10.7-21.1) was similar to the estimates found in blood donors in the Central Region of Argentina which includes Santa Fe (12.9%) (Flichman et al. 2022). However, it was lower than the seroprevalence of anti-DENV antibodies among adult individuals who reported no prior infection with dengue in Posadas (20.5%), a city located in northeastern Argentina [15]. This difference may be related to the persistence of detectable DENV antibodies throughout life, and the longer history of dengue epidemics in Posadas with outbreaks occurring more frequently since 2000. Therefore, the seroprevalence in Posadas is expected to be higher than in Santa Fe due to the increased probabilities of repeated exposure to the virus over time.

In our study, which included individuals over 18 years of age, we did not find any significant difference in seropositivity by age group. However, studies conducted in hyperendemic areas have found an association with age and attribute it to the fact that the longer people reside in a dengue-endemic region, the greater their risk of exposure, coupled with the lifelong persistence of anti-DENV antibodies [28,29]. In the city of Santa Fe, where the first severe dengue outbreak occurred in 2016 [5], all age groups studied have been exposed to the outbreaks that have occurred since then. Therefore, we can consider that the absence of variation by age in our study could be attributed to the relatively recent onset of dengue outbreaks in this region.

We were surprised to find that knowledge about the disease symptoms and modes of transmission was not associated with a lower probability of seropositivity. However, other studies have also found a similar pattern regarding knowledge, attitudes and practices (KAPs) and seropositivity to DENV, *Leptospira* spp. or SARS-COV 2 [30–33].

One of the strengths of our study is that it is based on data collected *in situ*, using a survey instrument and direct observation of the socio-environmental context. Contrary to what we expected, we found lower probabilities of DENV seropositivity in residents with neighboring vacant lots compared to those with neighboring houses. A number of studies have found greater abundance of *A. aegypti* larvae on vacant lots in comparison to houses, but they attribute this to the possibility that vacant lots with high vegetation cover can hold litter that offer breeding sites for mosquitoes [34–36]. At our study sites, small dump sites are more often found in backyards, whereas vacant lots mostly encompass green areas on the riverside or open fields in these less urbanized settings. Therefore, we interpret our finding as resulting from greater transmission of the virus occurring in more urban contexts with greater human density than in more rural or suburban settings [37]. *Aedes aegypti* is considered a highly anthropophilic vector that has adapted to live and breed in human dwellings [35]. Our results support this view and underscore the importance of targeting buildings and peridomiciliary areas during interventions aimed at controlling mosquitoes [38].

One limitation of this study was information bias related to the sensitivity of the ELISA performed. Given that the sensitivity may not be particularly high (84.8%), the real seroprevalence may be higher than what was found in the study. Conversely, seroprevalence could also be overestimated due to cross-reactivity with antibodies against other flaviviruses, typically detected using the plaque reduction neutralization test [39], an assay that was not part of this research. Another caveat worth noting is the potential source of bias in the estimated seroprevalence due to the group of eligible participants who agreed to complete the epidemiological survey but refused to provide a blood sample. This refusal was significantly higher in site CH (*p* = 0.003), the most urbanized neighborhood, where 41% of the selected participants did not provide a blood sample compared to 20-22% in the other two neighborhoods. Additionally, it should be noted that 44% of the people who did not agree to provide a blood sample were surveyed during January-March 2020, and the news about the emergence of COVID-19 may have made them reluctant to participate. The beginning of the national lockdown due to the pandemic, which overlapped with the 2020 dengue outbreak, also impeded us from continuing to collect blood samples and thus enhance the statistical power of the study.

Despite the aforementioned limitations, our study provides valuable insights into the risk factors for DENV seropositivity in residents of neighborhoods in Santa Fe, Argentina, and underscores the importance of population screening to obtain estimates of the number of cases that are not detected by the National Health Surveillance system. With the emergence of epidemic dengue and the more common occurrence of severe forms of DENV infections (DHF/DSS) it is critical to shed light into the factors that increase risk of infection to better manage this important public health challenge. Our findings can contribute to building consensus on the factors that increase DENV infection. Additionally, our findings will be valuable in designing public health interventions in vulnerable neighborhoods during dengue outbreaks.

## Data Availability

All data produced are available online at Mendicino, D., Ricardo, T., Cristaldi, M., Maglianesi, M., Guzman, G., Claussen, S., Chiaraviglio, R. G., AVALOS, C., & Previtali, M. A. (2024). Data for Seropositivity to Dengue virus DENV in three neighborhoods in the periphery of a city with a recent history of outbreaks in Argentina: what can we learn from unreported cases? (version1.0) [Data set]. Zenodo. https://doi.org/10.5281/zenodo.11507563

## Acknowledgements

We want to thank technicians that assisted in the field sampling: Leda Beltramo, Mercedes Gaggiamo, Martín Pinto, Gabriela Mertes. Additionally, we want to acknowledge the contribution to the survey questionnaire on Astor Borotto and Gabriel Obradovich. Finally, we are thankful to the neighbors that participated in the study and to social organizations that facilitated our outreach to the community: Garganta Poderosa, Biblioteca Popular de Las Orillas, La Revuelta.

## Declaration of Generative AI and AI-assisted technologies in the writing process

During the preparation of this work the author(s) used Chat GPT-3.5 in order to check the grammar and spelling of some parts of the manuscript. After using this tool/service, the author(s) reviewed and edited the content as needed and take(s) full responsibility for the content of the publication.

## Supplementary information

Supplementary File 1. STROBE Statement—Checklist of items that should be included in reports of cross-sectional studies

Supplementary File 2. Ricardo, T. (2024). Issengard83/xlsx2geojson: Version 1.0 (version1.0). Zenodo. https://doi.org/10.5281/zenodo.11549812

